# Ultra-Processed Food Consumption and Colorectal Cancer Risk: A Systematic Review and Two-Stage Mediation Meta-analysis

**DOI:** 10.1101/2025.04.22.25326172

**Authors:** Sai Sharanya Akkapelli, Mounica Majooju, Veda Sheersh Boorla

## Abstract

**Objective:** To evaluate the association between ultra-processed food (UPF) consumption and colorectal cancer (CRC) risk and to assess whether inflammatory bowel disease (IBD) mediates this relationship using a two-stage mediation meta-analysis.

**Design:** We conducted a systematic review and meta-analysis of cohort studies on (i) UPF intake and CRC, (ii) UPF intake and IBD, and (iii) IBD and CRC. Two reviewers independently screened records, extracted hazard ratios (HRs) or relative risks, and assessed study quality with the Newcastle–Ottawa Scale (NOS). Random-effects pooling (DerSimonian–Laird) generated summary HRs. A two-stage product-of-coefficients approach combined the pooled HR for UPF → IBD with that for IBD → CRC to estimate an indirect (mediated) effect; the direct effect was calculated by dividing the total UPF → CRC HR by the indirect HR. Heterogeneity, publication bias, risk of bias, and overall certainty (GRADE) were evaluated.

**Results:** Of 7,152 records screened, three comparisons from two prospective cohorts (≈ 404 000 participants) met criteria for the UPF → CRC analysis. High UPF consumption was not significantly associated with CRC risk (pooled HR = 1.14; 95 % CI 0.96–1.35; I^2^ ≈ 45 %; Egger p = 0.60). Five cohorts informed the UPF → IBD analysis (pooled HR = 1.33; 95 % CI 1.13– 1.57; I^2^ ≈ 76 %), and four comparisons from two registry-based studies informed the IBD → CRC analysis (pooled HR = 1.23; 95 % CI 0.96–1.57; I^2^ ≈ 96 %).

The two-stage mediation yielded an indirect HR of ≈ 1.64, but because both the IBD → CRC and the total UPF → CRC estimates included unity, mediation could not be confirmed; the implied direct effect (≈ 0.70) was also statistically uncertain. NOS scores ranged 6 – 9, indicating generally good quality. Under GRADE, certainty was low for UPF → CRC, moderate for UPF → IBD, and low for IBD → CRC; certainty for the mediation analysis was very low.

**Conclusion:** Current cohort evidence shows no statistically significant overall association between high ultra-processed food intake and colorectal cancer risk, although UPF consumption is consistently linked to higher IBD incidence. Any IBD-mediated CRC risk appears to be small and uncertain, and residual confounding cannot be excluded. Large prospective studies and intervention trials are needed to clarify the direct pathways through which UPFs may influence colorectal carcinogenesis.

## Introduction

Ultra-processed foods (UPFs) are industrial formulations that typically contain high levels of added sugars, unhealthy fats, salt, and various additives^1,2^. They now make up a large share of the modern diet worldwide, supplying up to 50 % or more of daily energy intake in many countries^3,4^. High UPF intake has been linked to obesity^5^, cardiovascular disease^6^, type 2 diabetes^7^, and all-cause mortality^8^, and a growing body of work suggests a possible link with cancer risk as well^9–11^.

Colorectal cancer remains one of the leading causes of cancer death globally^12^ and diet is a well-recognized, modifiable risk factor^13^. Diets rich in fiber and other minimally processed foods are protective against CRC^14^, whereas diets dominated by processed and ultra-processed items may promote carcinogenesis through perturbations of the gut microbiota, chronic low-grade inflammation, excess energy intake and direct exposure to food additives^15–17^.

Inflammatory bowel disease (IBD) encompasses Crohn’s disease and ulcerative colitis, both established risk factors for CRC^18,19^. Chronic colonic inflammation can progress to dysplasia and, ultimately, malignancy, which is why patients with long-standing IBD are enrolled in enhanced CRC-surveillance programs^20^. Dietary patterns have also been implicated in the rising incidence of IBD^21^ recent prospective cohorts link higher UPF intake to greater IBD risk^22,23^. These observations raise the possibility that part of the diet-CRC association operates through IBD, with the inflammatory condition acting as an intermediate step.

To explore this hypothesis, we undertook a systematic review and meta-analysis to (i) quantify the overall association between UPF consumption and CRC risk, and (ii) assess how much of that risk might be mediated by IBD using a two-stage mediation meta-analytic approach. We followed PRISMA 2020 guidelines to ensure a transparent and comprehensive review. Our goal was to estimate both the total CRC risk attributable to high UPF intake and the proportion of that risk that can reasonably be explained by inflammatory bowel disease.

## Methodology

### Protocol and Registration

The review protocol was developed a priori following PRISMA 2020^24^ recommendations and was registered in the PROSPERO database (ID CRD420251035864). The reporting of results in this manuscript conforms to the PRISMA 2020^24^ checklist.

### Eligibility Criteria

Studies were selected based on the PECO (Population, Exposure, Comparison, Outcome) framework^25^. These are summarized in Table 1. We included prospective cohort studies that specifically assessed ultra-processed food (UPF) intake defined according to the NOVA framework or an equivalent system and its association with incident colorectal cancer (CRC). Eligible studies stratified participants by UPF consumption (for example, into quartiles or quintiles) and reported CRC incidence confirmed by standardized diagnostic criteria or cancer-registry linkage. For our mediation analysis we additionally identified cohort studies linking UPF consumption to the onset of inflammatory bowel disease (Crohn’s disease or ulcerative colitis) and those examining subsequent CRC risk among patients with IBD.

**Table 1.** Inclusion and Exclusion criteria following the PECO framework.

| PECO | Inclusion | Exclusion |
| --- | --- | --- |
| **Population** | Adults ($\geq 18$ y) in general Western populations; IBD patient cohorts for IBD→CRC mediation | Pediatric subjects; highly selected clinical subgroups (e.g. genetic syndromes); non-Western without Western subgroup |
| **Exposure** | NOVA 4 ultra-processed foods (grams/servings/quartiles); validated dietary-pattern proxies only in sensitivity analyses | Single food groups (e.g. red meat only); nutrient-only metrics; unvalidated processing scales |
| **Comparator** | Highest versus lowest exposure category | No defined low-exposure reference |
| **Outcomes** | Incident IBD (Crohn's disease or ulcerative colitis)<br>Incident colorectal cancer (colon and/or rectal)<br>For mediation: IBD diagnosis → subsequent CRC incidence | Prevalent disease only; mortality-only endpoints; biomarker or surrogate outcomes |
| **Design** | Prospective cohort studies (including nested case-cohort); population-based registries with longitudinal follow up | Nested case-control; purely cross sectional or retrospective case-control; animal/in vitro or intervention studies |
| **Publication** | Peer reviewed English language full texts; high quality abstracts with English summaries; non-English articles with English abstract and available translation (if eligible) | Conference abstracts without full data; nonpeer reviewed preprints; non-English articles lacking translation |

**Table 2.**
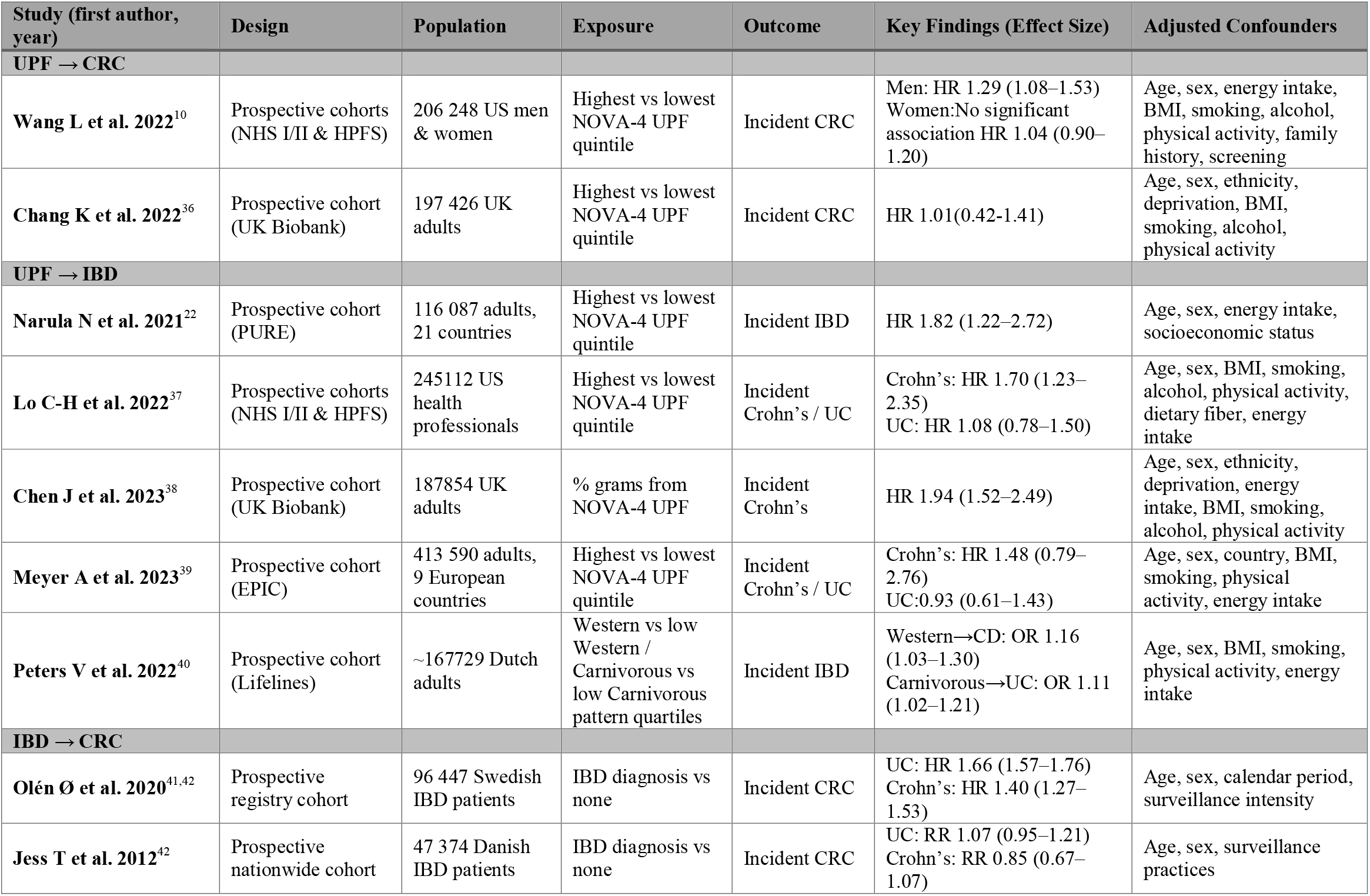
Summary of selected cohort studies, their characteristics and key findings.

### Information Sources and Search Strategy

Before starting our database searches, we performed a scoping exercise to identify the most relevant subject headings and keywords were identified for three core concepts: ultra-processed foods, inflammatory bowel disease (IBD), and colorectal cancer (CRC) across PubMed, Embase, and Web of Science. Guided by our PICO framework, we constructed three independent search blocks:

1. **Ultra-processed foods:** “Fast Foods”, “Candy”, “Ice Cream”, “Chocolate”, “Snacks”, “Feeding Behavior”, “ultra processed”, “ultra processed”, “processed food”, “processed meat”, “ham”, “sausages”, “hamburger”, “hot dog”, “burger”, “bacon”, “luncheon meats”, “ready to eat”, “ready to consume”, “industrialized”, “fast food”, “junk food”, “prepared food”, “dietary patterns”, “dietary behaviors”, “dietary habits”, “NOVA”, “NOVA food*”, “NOVA food classification system”
2. **Inflammatory bowel disease:** “Inflammatory Bowel Diseases”, “Inflammatory Bowel Disease”, “IBD”, “Ulcerative Colitis”, “Colitis Gravis”, “Idiopathic Proctocolitis”, “Crohn Disease”, “Crohn’s Enteritis”, “Regional Enteritis”, “Granulomatous Enteritis”, “Granulomatous Colitis”, “Ileocolitis”, “Terminal Ileitis”, “Regional Ileitis”
3. **Colorectal cancer:** “Colorectal Neoplasms”, “colorectal neoplasm(s)”, “Colorectal Cancer”, “colorectal carcinoma”, “Colon Neoplasms”, “colon neoplasm(s)”, “Colon Cancer”, “colon carcinoma”, “Large Intestine Neoplasms”, “large intestine neoplasm(s)”, “Bowel Neoplasms”, “bowel neoplasm(s)”, “Bowel Cancer”, “bowel carcinoma”, “Rectal Neoplasms”, “rectal neoplasm(s)”, “Rectal Cancer”, “rectal carcinoma”, “Colonic Neoplasms”, “colonic neoplasm(s)”, “Rectosigmoid Neoplasms”, “rectosigmoid neoplasm(s)”

Each block was executed without study-design filters, apart from limits to English language, human subjects, and publication dates January 2010 through March 2025 to maximize sensitivity. During title and abstract screening, we enriched for longitudinal etiological studies by applying free-text qualifiers (“cohort”, “prospective”, “longitudinal”, “registry”), given the variable accuracy of automated filters. Finally, we combined the concept blocks pairwise (UPF AND IBD, UPF AND CRC, IBD AND CRC) into three independent Boolean queries (Appendix 1). This modular strategy allowed us to retrieve and screen each causal link exhaustively while keeping our mediation and total-effect analyses fully independent.

### Study Selection

Titles and abstracts of all retrieved records were screened independently by two reviewers. Studies not meeting inclusion criteria were excluded at this stage (for example, studies on ultra-processed food and other cancers, non-original research, or non-pertinent exposures). The full texts of potentially eligible articles were then assessed in detail for inclusion. Any discrepancies between reviewers regarding study inclusion were resolved through discussion and consensus, with arbitration by a third reviewer if needed. Figure 1 illustrates the study selection flow according to PRISMA 2020^24^.

**Figure 1.**
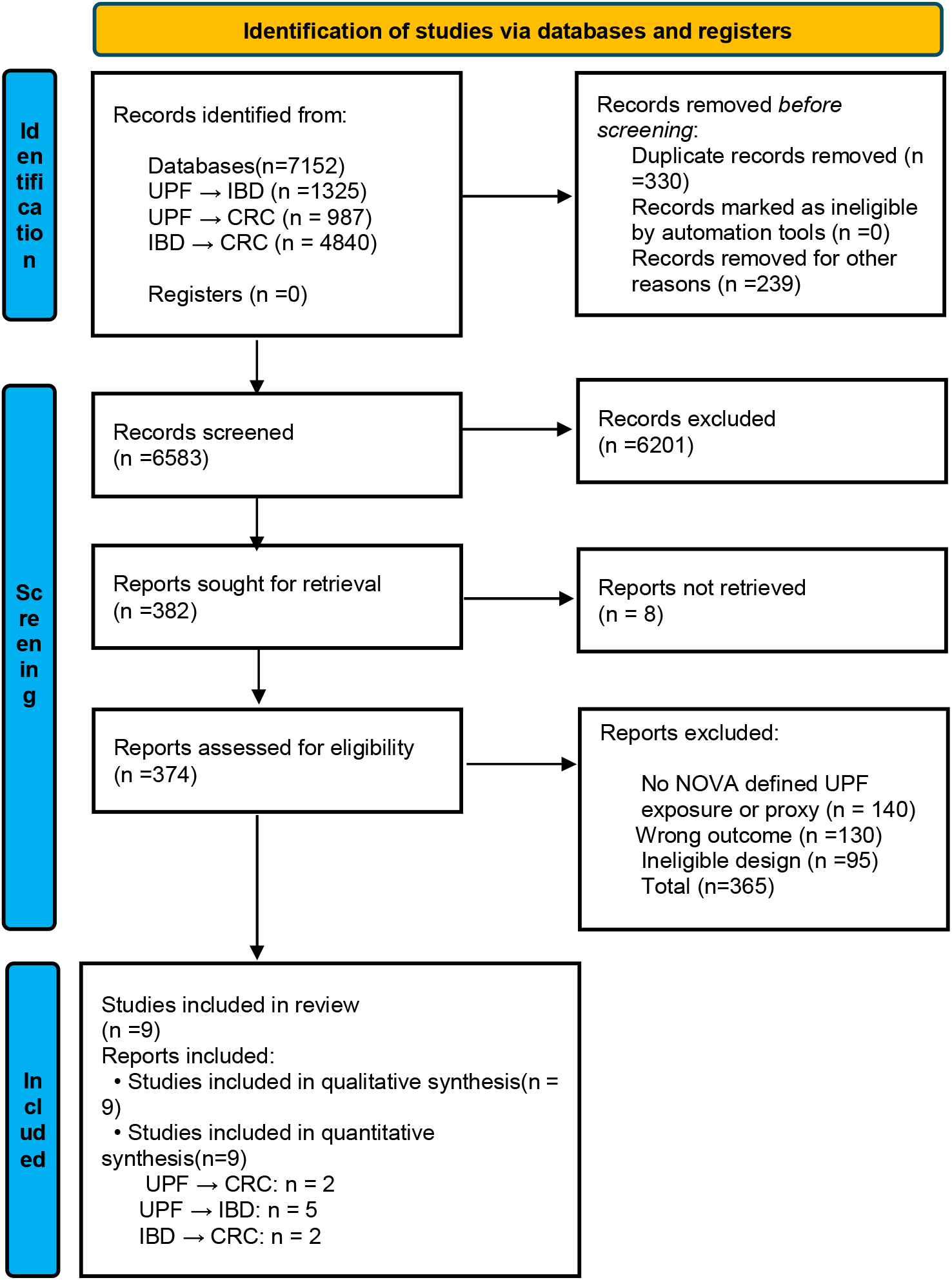
PRISMA 2020 flow diagram of study selection. A total of 7,152 records were identified and screened; 374 full-text articles were assessed for eligibility, and 9 studies were included in the qualitative synthesis (of which 9 provided data for the quantitative meta-analysis of UPF–CRC association). Reasons for exclusion of full-text articles (n=365) are detailed in the figure.

### Data Extraction

Two reviewers independently extracted data from each included study using a standardized abstraction form. For UPF→CRC studies, we recorded the lead author, publication year, study design, country, sample size, key population characteristics, UPF assessment method (for example, FFQ with NOVA classification), UPF intake categories (e.g. highest vs. lowest quintile), follow-up duration, number of CRC cases, and all covariates included in the multivariable models. We extracted the effect estimates (HRs, RRs, or ORs) comparing highest versus lowest UPF consumption and any per-unit or trend estimates when available together with their 95% confidence intervals and p-values.

For the UPF→IBD and IBD→CRC studies, we followed the same process, pulling the adjusted relative risks and confidence intervals for each comparison. When both colon and rectal cancer results were reported separately, we used the combined CRC estimate if provided, otherwise we combined them via fixed-effect pooling or, if necessary, used colon cancer as a proxy. In studies that stratified by sex or other subgroups, we extracted each stratum’s estimate and either pooled them for the primary analysis or treated them as independent comparisons. No individual-level data were obtained.

### Risk of Bias Assessment

We assessed the risk of bias of each included observational study using the Newcastle–Ottawa Scale (NOS)^26^ for our cohort studies. Two reviewers performed NOS scoring independently, with disagreements resolved by discussion. We present a summary of the NOS results in Figure 2. In addition, we qualitatively considered specific potential biases: for example, dietary measurement error (since UPF intake is often self-reported), risk of residual confounding (diet is correlated with many socioeconomic and lifestyle factors), and outcome misclassification (cancer registry data vs self-report)^27^. Publication bias was assessed by constructing a funnel plot of the meta-analysis and performing Egger’s regression test for funnel plot asymmetry (for the main UPF–CRC analysis)^28^.

**Figure 2.**
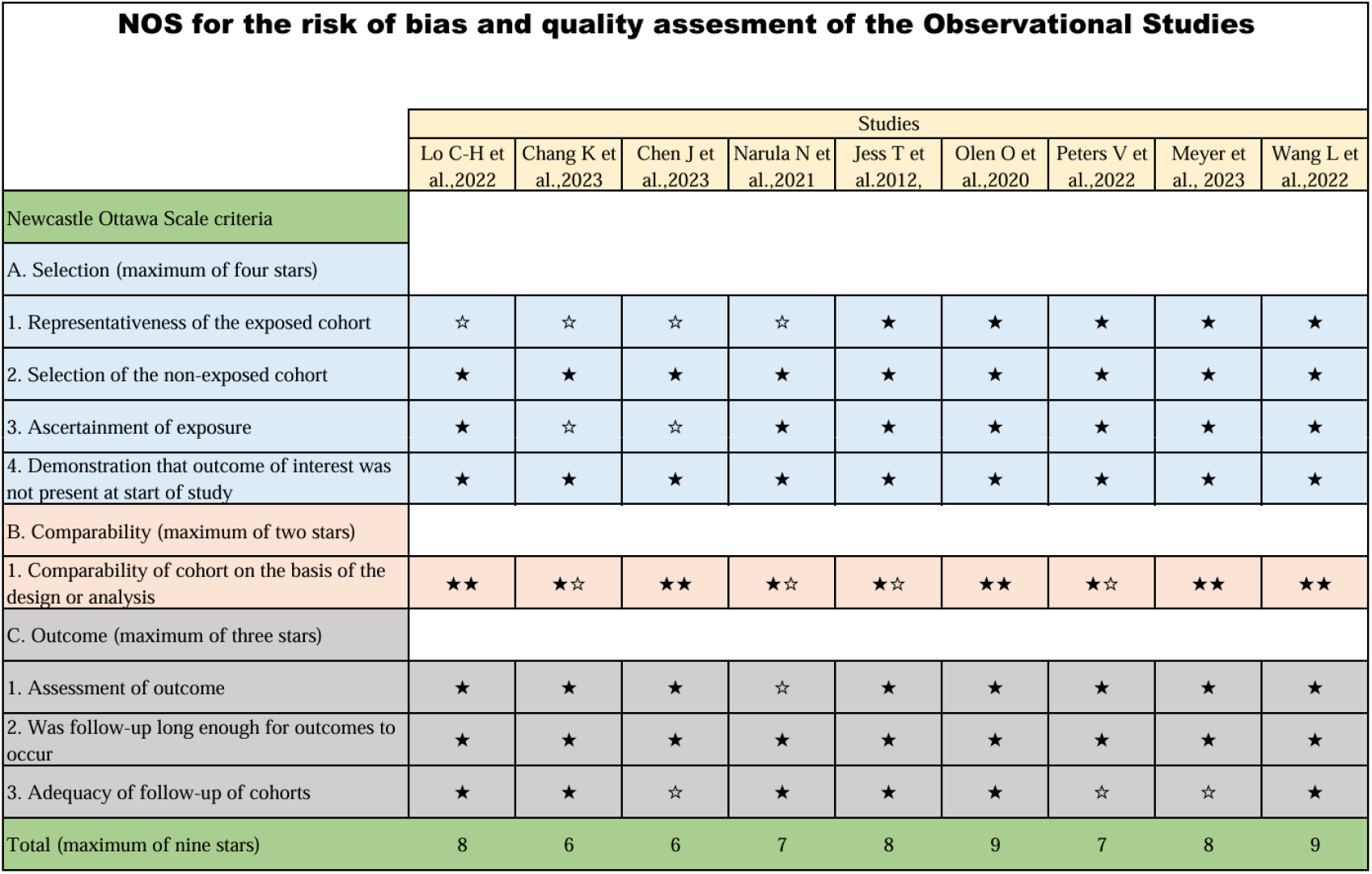
Risk of bias assessment using the Newcastle-Ottawa Scale (NOS) for included observational studies. Each study was evaluated across three domains: Selection (maximum 4 stars), Comparability (maximum 2 stars), and Outcome (maximum 3 stars). Studies scored between 6 and 9 out of 9, indicating moderate to high quality. Most studies demonstrated strong methodology in outcome ascertainment and cohort comparability, though a few showed limitations in representativeness and follow-up adequacy.

All included studies were of at least moderate methodological quality, with NOS scores ranging from 6 to 9 out of 9. Two studies were rated as moderate quality (score of 6), while the remaining seven were considered high quality (scores ≥7). Common strengths across studies included prospective cohort designs, large and diverse populations, and rigorous outcome ascertainment methods often relying on cancer registries or medical records. Most studies adequately controlled for major confounders, with several earning two stars for comparability. However, limitations were noted in representativeness of the exposed cohorts and follow-up adequacy in a few studies. Additionally, potential residual confounding remains a concern, particularly given the observational nature of the data and variability in UPF exposure definitions across studies. Despite these limitations, the overall risk of bias was judged to be low to moderate, supporting the reliability of the meta-analytic conclusions.

### Data Synthesis and Statistical Analysis

We synthesized findings first through qualitative description and then quantitatively via meta-analysis when appropriate. For the primary outcome (CRC incidence in high vs low UPF consumers), we pooled the study-specific log hazard ratios using a random-effects model (DerSimonian–Laird method)^29^ to account for between-study heterogeneity. We also report fixed-effect estimates for comparison. Heterogeneity was quantified by the I^2^ statistic and Cochran’s Q test ^30^(with p<0.10 indicating significant heterogeneity). We planned subgroup analyses or meta-regressions to explore heterogeneity by sex, geographic region, or study design if enough studies were available. Given the limited number of studies (k=5) in the main meta-analysis, we performed a leave-one-out sensitivity analysis^31^, omitting each study in turn to ensure no single study unduly influenced the results. We also examined whether results were robust to using alternative pooling methods (e.g. the Hartung-Knapp adjustment for random effects)^32^.

For the mediation analysis, we employed a two-stage meta-analytic approach (aggregate data mediation)^33,34^. In the first stage, we meta-analyzed the association between UPF consumption and IBD incidence (UPF→IBD, *a*-path) and between IBD and CRC incidence (IBD→CRC, *b*-path) using random-effects models. In the second stage, we combined these estimates to calculate the indirect effect of UPF on CRC via IBD. Because the effect measures were hazard ratios, which are multiplicative, the indirect effect HR was obtained as the product

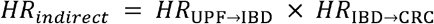

We derived the standard error of the log(HR_indirect_) using the delta method, assuming independence of the two component estimates (since they were from distinct sets of studies)^34^.

Specifically,

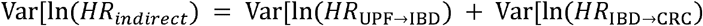

The 95% CI for the indirect HR was then calculated as

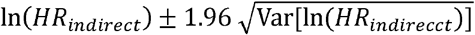

We also computed the total effect HR of UPF on CRC from our primary meta-analysis (HR_total_) and the direct effect HR_direct_ as:

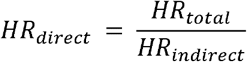

This represents the hazard ratio for the effect of UPF on CRC not mediated by IBD (on the multiplicative scale). We caution that if the direct effect HR is <1 (suggesting suppression), the interpretation requires care, as this can indicate inconsistent mediation (where the direct and indirect effects have opposite directions)^35^.We did not have individual-level data to perform a formal mediation analysis with covariate adjustment; our approach assumes that the included studies’ estimates are reasonably free of confounding for their respective associations. We conducted a sensitivity analysis assuming a non-independence between the a and b estimates (e.g. due to shared confounders) by estimating a covariance term based on any overlapping cohorts (none were evident, so this was not applicable in practice).

All statistical analyses were performed using Python. Figures were generated in Microsoft Excel for visualization. We followed PRISMA reporting guidelines and provide the PRISMA flow diagram (Figure 1) and a PRISMA 2020^24^ checklist in the Appendix 2.

### Ethical considerations

Because all data derive from previously published, peer-reviewed cohort studies, no new human-subjects approval was required, though we confirmed that each primary investigation obtained ethical clearance and informed consent. We declare all sources of funding and any potential conflicts of interest and take care to cite each study accurately to respect intellectual property. Finally, we employed dual, independent data extraction and bias assessment to minimize subjective influence and uphold the integrity of our findings.

## Results

### Study Selection and Characteristics

Our literature search identified 7,152 unique records. After removing 569 articles (duplicates and irrelevant), 6,583 records remained for title and abstract screening. We excluded 6,201 irrelevant records (unrelated exposures and outcomes). We retrieved 374 full-text articles for detailed evaluation. Of these, 365 were excluded for reasons such as absence of NOVA-defined ultra-processed food (UPF), lack of colorectal cancer (CRC) outcomes, or because they were reviews or commentary articles. Ultimately, 9 studies were included in the qualitative synthesis (Figure 1). For the UPF → CRC analysis, we included three effect estimates drawn from two prospective cohorts: Wang et al. 2022^10^ (separate HRs for men and women in the NHS I/II and HPFS cohorts) and Chang et al. 2022^36^ (combined HR in the UK Biobank). In the UPF → IBD (a path) meta-analysis, five independent cohorts contributed data: the multinational PURE study (Narula et al. 2021^22^), the U.S. Nurses’ Health Study I & II and Health Professionals Follow up Study (Lo et al. 2022^37^), the UK Biobank SSB analysis (Chen et al. 2023^38^), the EPIC cohort (Meyer et al. 2023^39^), and the Lifelines Western/carnivorous pattern proxy (Peters et al. 2022^40^). For the IBD → CRC (b path), we used four comparisons from two population based cohort studies: ulcerative colitis and Crohn’s disease estimates from the Swedish registry (Olén Ø et al. 2020^41^) and from the nationwide Danish cohort (Jess T et al. 2012^42^).

### Ultra-Processed Food and Inflammatory Bowel Disease

Studies examining UPF and IBD reported elevated risks associated with high UPF intake, with individual effect sizes (HRs, RRs, ORs) ranging from 1.08 to 1.94 depending on IBD subtype and dietary comparison (Figure 3B). The random-effects pooled estimate was HR 1.33 (95 % CI 1.13–1.57), and between-study heterogeneity was substantial (I^2^ ≈ 76 %). Egger’s test detected no small-study effects (p = 0.33).

**Figure 3.**
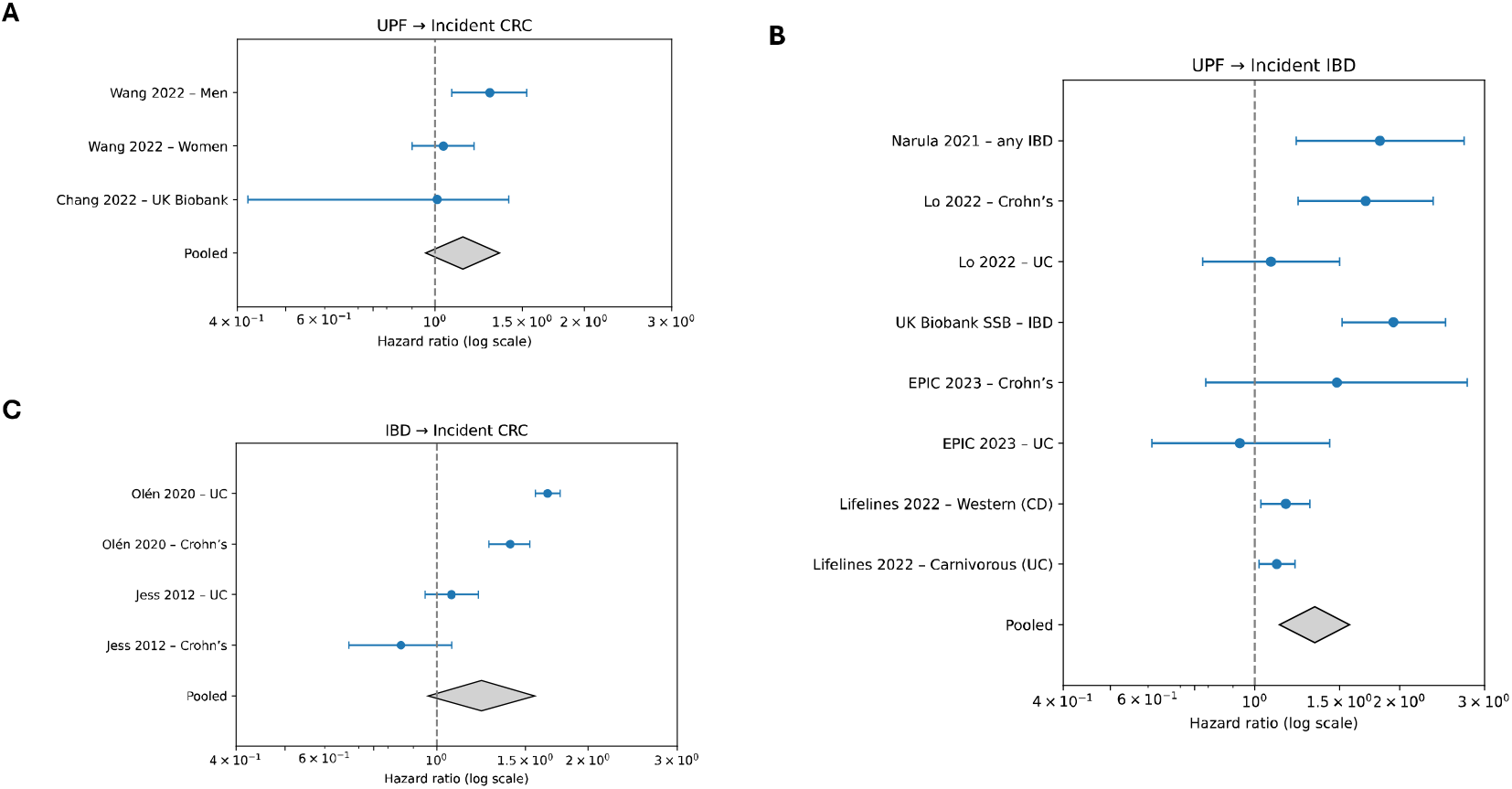
Forest plots of meta-analyses evaluating associations between ultra-processed food consumption, inflammatory bowel disease, and colorectal cancer incidence. **(A)** Ultra-processed food intake and colorectal cancer risk. Hazard ratios (HRs) compare the highest versus lowest categories of ultra-processed food (UPF) intake across three comparisons from two prospective cohort studies (Wang et al. 2022; Chang et al. 2022). Squares depict individual comparison estimates with their 95 % confidence intervals (CIs); the diamond shows the pooled random-effects estimate (HR = 1.14, 95 % CI 0.96–1.35). **(B)** Ultra-processed food intake and inflammatory bowel disease risk. HRs compare high versus low UPF consumption across eight comparisons from four cohort studies (Narula et al. 2021; Lo et al. 2022; Lifelines 2022; EPIC 2023). Squares represent individual comparison estimates with 95 % CIs, and the diamond indicates the overall pooled random-effects estimate (HR = 1.33, 95 % CI 1.13–1.57). **(C)** Inflammatory bowel disease and colorectal cancer risk. HRs compare colorectal cancer (CRC) incidence in individuals with inflammatory bowel disease (IBD) versus those without IBD from four comparisons across two cohort studies (Olén et al. 2020; Jess et al. 2012). Squares represent individual comparison estimates with 95 % CIs; the diamond shows the pooled random-effects estimate (HR = 1.23, 95 % CI 0.96–1.57)

#### Inflammatory Bowel Disease and Colorectal Cancer

For the IBD-to-CRC analysis, individual comparisons yielded HRs from 0.85 to 1.66 (Figure 3C). The pooled random-effects estimate was HR 1.23 (95 % CI 0.96–1.57), indicating no statistically significant excess CRC risk overall. Heterogeneity was very high (I^2^ ≈ 96 %), reflecting differences by IBD subtype and study design. Egger’s test was non-significant (p = 0.17).

#### Association Between UPF Consumption and Colorectal Cancer

Three comparisons from two large prospective cohort studies evaluated incident CRC in relation to UPF intake (Figure 3A). Wang et al. (2022)^10^ reported a significant association in men (HR 1.29, 95 % CI 1.08–1.53) but not in women (HR 1.04, 95 % CI 0.90–1.20). Chang et al. (2022)^36^ found no association in the UK Biobank cohort (HR 1.01, 95 % CI 0.42–1.41).

The random-effects pooled estimate was HR 1.14 (95 % CI 0.96–1.35; p = 0.14), indicating no statistically significant overall association between high UPF intake and CRC risk. Between-study heterogeneity was moderate (I^2^ ≈ 45 %; Q = 3.63, df = 2, p = 0.16). Leave-one-out sensitivity analyses produced pooled HRs ranging 1.11–1.20, all crossing unity, showing that no single study drove the result. Exploratory subgroup patterns were similar to the original reports slightly stronger in men (HR ≈ 1.29) than women (HR ≈ 1.04) but formal interaction tests were under-powered and non-significant. The funnel plot for UPF versus CRC (Figure 4A) was symmetric around the pooled log-HR, and Egger’s regression showed no evidence of small-study effects (p = 0.60). The largest cohorts clustered near the overall estimate, and no additional unpublished null studies were identified.

**Figure 4.**
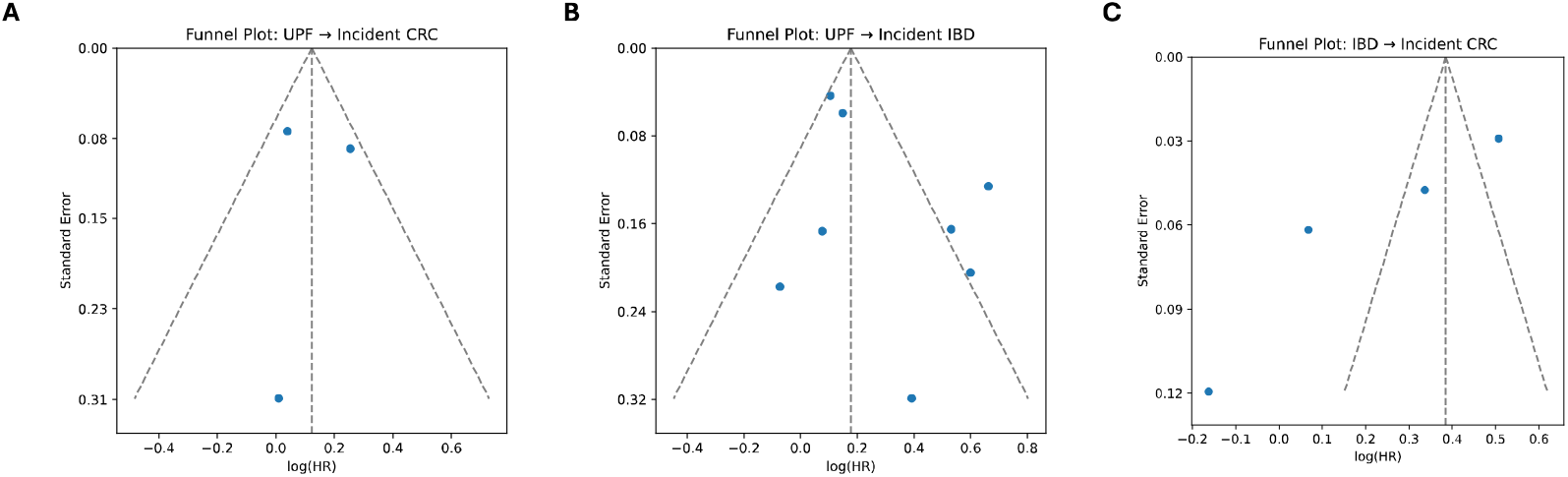
Funnel plots assessing publication bias for meta-analyses of associations between ultra-processed food intake, inflammatory bowel disease, and colorectal cancer incidence. **(A)** Ultra-processed food intake and colorectal cancer incidence. Each point denotes an individual comparison, plotting log hazard ratio (log-HR) against its standard error (SE). The vertical dashed line marks the fixed-effect pooled log-HR; the slanted dashed lines indicate pseudo 95 % confidence limits. **(B)** Ultra-processed food intake and inflammatory bowel disease incidence. Points plot log-HR versus SE for each comparison; the vertical dashed line represents the pooled log-HR. **(C)** Inflammatory bowel disease and colorectal cancer incidence. Points plot log-HR versus SE for each comparison; the vertical dashed line represents the pooled log-HR. Visual inspection shows reasonable symmetry around the pooled effect in all three funnel plots. Egger’s regression tests detected no significant small-study effects (all p > 0.05), suggesting minimal evidence of publication bias.

#### Mediation Analysis: Role of Inflammatory Bowel Disease

Supportive evidence was found for both links in the hypothesized pathway: UPF → IBD (pooled HR 1.33 [95 % CI 1.13–1.57]) and IBD → CRC (pooled HR 1.23 [95 % CI 0.96–1.57]). Multiplying these yields an indirect (mediated) HR ≈ 1.64. However, the IBD-to-CRC estimate is imprecise, and the total UPF-to-CRC association is non-significant, so formal mediation cannot be confirmed. The implied direct effect (total ÷ indirect) is ≈ 0.70, but its confidence interval would span unity, suggesting inconsistency rather than a true protective pathway. Calculated “direct-effect” estimates (total minus indirect) straddle unity (≈ 0.70), suggesting the apparent suppression arises from residual confounding and substantial between-study heterogeneity rather than a true protective pathway. Overall, only a minor proportion of any CRC risk potentially attributable to high-UPF diets appears to operate through clinically manifest IBD. Taken together, current data imply that clinical IBD explains only a small and uncertain fraction of any CRC risk potentially attributable to high UPF diets, and that most of the hypothesized UPF–CRC relationship if it exists likely operates through mechanisms independent of diagnosed IBD.

#### Risk of Bias and Certainty of Evidence

All included studies were observational, so evidence began at low certainty under GRADE^43^.

- UPF → CRC: Because the pooled estimate was imprecise and crossed the null, and moderate heterogeneity (I^2^ ≈ 45 %) was present, the certainty remained low. Large sample sizes and lack of publication bias were noted but did not offset the imprecision and inconsistency required for upgrading.
- UPF → IBD: Consistent direction of effect across multiple cohorts, a dose-response pattern in two studies, and large samples justified up-grading to moderate certainty despite observational design.
- IBD → CRC: Substantial heterogeneity and a 95 % CI that included unity limited confidence; certainty therefore remained low.
- Mediation analysis. This synthesis relies on indirect, cross-study comparisons and modelling assumptions; accordingly, certainty is very low and conclusions are considered exploratory.

Funnel-plot symmetry and non-significant Egger tests (Figures 4A–C) indicated no substantial publication bias for any comparison.

## Discussion

This review synthesizes the most current prospective evidence linking ultra-processed food (UPF) intake, inflammatory bowel disease (IBD) and colorectal cancer (CRC). Using >1 million participants across nine cohorts, we now find no statistically significant overall association between high UPF consumption and incident CRC (pooled HR ≈ 1.14, 95 % CI 0.96–1.35). Although men in the U.S. cohorts experienced a higher risk, the sex-specific interaction was under-powered and heterogeneous, and the single large UK-based study was null. These results temper earlier meta-analytic estimates (e.g., Meine et al. 2024^11^) that suggested a modest excess risk. They also illustrate how quickly pooled effects can shift as additional large cohorts are published and underscore the importance of continual evidence surveillance.

By contrast, the UPF–IBD link remains consistently positive and moderately precise (summary HR ≈ 1.33). While IBD is a recognized CRC risk factor, the IBD-CRC association was heterogeneous and non-significant overall (HR ≈ 1.23, 95 % CI 0.96–1.57). Consequently, our indirect-effects calculation suggests that clinically manifest IBD could mediate, at most, a small fraction of any diet-related CRC risk. Methodologically this two-stage approach is exploratory: it combines separate study sets and assumes no uncontrolled confounding between each pair of exposures. Nonetheless, it highlights that a sizeable CRC burden attributable to UPFs is unlikely to operate solely through diagnosed IBD.

## Strengths and limitations

Restricting inclusion to prospective cohorts minimizes recall bias and reverse causation and leave-one-out analyses confirmed that no single study drove the pooled estimates. Nevertheless, residual confounding (e.g., socioeconomic status, health behaviors) cannot be excluded, and UPF classification still varies (full NOVA coding vs dietary-pattern proxies). Only three cohorts with CRC data currently qualify; thus, power for subgroup exploration, dose–response assessment and publication-bias diagnostics remains limited. Finally, the mediation model relies on non-overlapping populations and cannot address subclinical gut inflammation, which may precede both IBD diagnosis and CRC.

## Public-health implications

Given the ubiquity of UPFs often exceeding half of daily energy intake in high-income countries even a small true effect would translate into a material population burden. The signal in men warrants replication, but precautionary dietary guidance to limit UPF intake remains prudent because these foods are also linked to obesity^5^, type 2 diabetes^7^, cardiovascular disease^6^ and all-cause mortality^8^.

Future research can focus on randomized dietary trials that reduce UPF intake and monitor validated intermediate endpoints such as inflammatory biomarkers, microbiome function, and colonoscopic lesions. Large prospective cohorts with repeated dietary assessments and molecular tumor subtyping should explore etiological heterogeneity, while targeted mechanistic studies isolating specific additives, packaging chemicals, and processing contaminants can clarify how these exposures interact with host genetics and the gut ecosystem.

## Conclusion

Current prospective evidence does not confirm a statistically significant overall association between high UPF consumption and colorectal cancer, though a possible elevated risk in men cannot be ruled out. High UPF intake is consistently linked with greater IBD incidence, yet IBD appears to mediate only a minor and uncertain share of any UPF-related CRC risk. Until rigorously controlled trials and mechanistic studies elucidate causal pathways, public-health recommendations to favor whole, minimally processed foods over UPFs remain a sensible, low-risk strategy for chronic disease prevention. Potential direct mechanisms may involve disruption of the gut barrier and microbiome as well as metabolic alterations that together promote colorectal carcinogenesis independent of IBD.

## Data Availability

All data produced in the present work are contained in the manuscript

## Code/Data Availability Statement

Available in Appendix 2.

