## Appendix for "Ultra-Processed Food Consumption and Colorectal Cancer Risk: A Systematic Review and Two-Stage Mediation Meta-analysis"

**Supplementary Appendix 1  Database Search Strategies**

*(executed 15 – 20 March 2025; Articles from 2010; English and Human filters applied)*

**Table 1. Ultra‑processed Food → Inflammatory Bowel Disease (UPF → IBD)**

| **Database** | **Search string** |
| --- | --- |
| **PubMed/MEDLINE** | "Fast Foods"[MeSH Terms] OR "Candy"[MeSH Terms] OR "Ice Cream"[MeSH Terms] OR "Chocolate"[MeSH Terms] OR "Snacks"[MeSH Terms] OR "Feeding Behavior"[MeSH Terms] OR "ultra-processed"[Title/Abstract] OR "ultraprocessed"[Title/Abstract] OR "Fast Foods"[Title/Abstract] OR "processed food"[Title/Abstract] OR "ultraprocessed food"[Title/Abstract] OR "ultra-processed food"[Title/Abstract] OR "processed meat"[Title/Abstract] OR "ultra-processed food"[Title/Abstract] OR "ham"[Title/Abstract] OR "sausages"[Title/Abstract] OR "hamburger"[Title/Abstract] OR "bacon"[Title/Abstract] OR "luncheon meats"[Title/Abstract] OR "ready-to-eat"[Title/Abstract] OR "ready-to-consume"[Title/Abstract] OR "industrialized"[Title/Abstract] OR "fast-food"[Title/Abstract] OR "fast-food"[Title/Abstract] OR "fast-food"[Title/Abstract] OR "junk food"[Title/Abstract] OR "prepared food"[Title/Abstract] OR "Candy"[Title/Abstract] OR "Ice Cream"[Title/Abstract] OR "Chocolate"[Title/Abstract] OR "Snacks"[Title/Abstract] OR "hot dog"[Title/Abstract] OR "burger"[Title/Abstract] OR "dietary patterns"[Title/Abstract] OR "dietary behaviors"[Title/Abstract] OR "dietary habits"[Title/Abstract] OR "NOVA"[Title/Abstract] OR "nova food*"[Title/Abstract] OR "NOVA food classification system"[Title/Abstract]  AND  ("Inflammatory Bowel Diseases"[MeSH Terms] OR "Inflammatory Bowel Diseases"[Title/Abstract] OR "Inflammatory Bowel Disease"[Title/Abstract] OR "IBD"[Title/Abstract] OR "colitis, ulcerative"[MeSH Terms] OR "colitis ulcerative"[Title/Abstract] OR "Idiopathic Proctocolitis"[Title/Abstract] OR "Ulcerative Colitis"[Title/Abstract] OR "Colitis Gravis"[Title/Abstract] OR "Crohn Disease"[MeSH Terms] OR "Crohn Disease"[Title/Abstract] OR "Crohn's Enteritis"[Title/Abstract] OR "Regional Enteritis"[Title/Abstract] OR "Crohn's Disease"[Title/Abstract] OR "Crohns Disease"[Title/Abstract] OR "Granulomatous Enteritis"[Title/Abstract] OR "Ileocolitis"[Title/Abstract] OR "Granulomatous Colitis"[Title/Abstract] OR "Terminal Ileitis"[Title/Abstract] OR "Regional Ileitis"[Title/Abstract]) |

**Table 2. Ultra‑processed Food → Colorectal Cancer (UPF → CRC)**

| **Database** | **Search string** |
| --- | --- |
| **PubMed/MEDLINE** | "Fast Foods"[MeSH Terms] OR "Candy"[MeSH Terms] OR "Ice Cream"[MeSH Terms] OR "Chocolate"[MeSH Terms] OR "Snacks"[MeSH Terms] OR "Feeding Behavior"[MeSH Terms] OR "ultra-processed"[Title/Abstract] OR "ultraprocessed"[Title/Abstract] OR "Fast Foods"[Title/Abstract] OR "processed food"[Title/Abstract] OR "ultraprocessed food"[Title/Abstract] OR "ultra-processed food"[Title/Abstract] OR "processed meat"[Title/Abstract] OR "ultra-processed food"[Title/Abstract] OR "ham"[Title/Abstract] OR "sausages"[Title/Abstract] OR "hamburger"[Title/Abstract] OR "bacon"[Title/Abstract] OR "luncheon meats"[Title/Abstract] OR "ready-to-eat"[Title/Abstract] OR "ready-to-consume"[Title/Abstract] OR "industrialized"[Title/Abstract] OR "fast-food"[Title/Abstract] OR "fast-food"[Title/Abstract] OR "fast-food"[Title/Abstract] OR "junk food"[Title/Abstract] OR "prepared food"[Title/Abstract] OR "Candy"[Title/Abstract] OR "Ice Cream"[Title/Abstract] OR "Chocolate"[Title/Abstract] OR "Snacks"[Title/Abstract] OR "hot dog"[Title/Abstract] OR "burger"[Title/Abstract] OR "dietary patterns"[Title/Abstract] OR "dietary behaviors"[Title/Abstract] OR "dietary habits"[Title/Abstract] OR "NOVA"[Title/Abstract] OR "nova food*"[Title/Abstract] OR "NOVA food classification system"[Title/Abstract]  AND  ("Colorectal Neoplasms"[MeSH Terms] OR "Colorectal Neoplasms"[Title/Abstract] OR "colorectal neoplasm"[Title/Abstract] OR "colorectal neoplasms"[Title/Abstract] OR "Colorectal Cancer"[Title/Abstract]  OR "colorectal carcinoma"[Title/Abstract] OR "Colon Neoplasms"[MeSH Terms] OR "Colon Neoplasms"[Title/Abstract] OR "colon neoplasm"[Title/Abstract] OR "colon neoplasms"[Title/Abstract]  OR "Colon Cancer"[Title/Abstract] OR "colon carcinoma"[Title/Abstract] OR "Large Intestine Neoplasms"[MeSH Terms] OR "Large Intestine Neoplasms"[Title/Abstract] OR "large intestine neoplasm"[Title/Abstract] OR "large intestine neoplasms"[Title/Abstract] OR "Bowel Neoplasms"[MeSH Terms] OR "Bowel Neoplasms"[Title/Abstract] OR "bowel neoplasm"[Title/Abstract] OR "bowel neoplasms"[Title/Abstract] OR "Bowel Cancer"[Title/Abstract] OR "bowel carcinoma"[Title/Abstract]  OR "Rectal Neoplasms"[MeSH Terms] OR "Rectal Neoplasms"[Title/Abstract] OR "rectal neoplasm"[Title/Abstract] OR "rectal neoplasms"[Title/Abstract] OR "Rectal Cancer"[Title/Abstract] OR "rectal carcinoma"[Title/Abstract] OR "Colonic Neoplasms"[MeSH Terms] OR "Colonic Neoplasms"[Title/Abstract] OR "colonic neoplasm"[Title/Abstract] OR "colonic neoplasms"[Title/Abstract] OR "Rectosigmoid Neoplasms"[MeSH Terms] OR "Rectosigmoid Neoplasms"[Title/Abstract] OR "rectosigmoid neoplasm"[Title/Abstract] OR "rectosigmoid neoplasms"[Title/Abstract]) |

**Table 3**. **Inflammatory Bowel Disease** **→ Colorectal Cancer** **(IBD → CRC)**

| **Database** | **Search string** |
| --- | --- |
| **PubMed/MEDLINE** | ("Inflammatory Bowel Diseases"[MeSH Terms] OR "Inflammatory Bowel Diseases"[Title/Abstract] OR "Inflammatory Bowel Disease"[Title/Abstract] OR "IBD"[Title/Abstract] OR "colitis, ulcerative"[MeSH Terms] OR "colitis ulcerative"[Title/Abstract] OR "Idiopathic Proctocolitis"[Title/Abstract] OR "Ulcerative Colitis"[Title/Abstract] OR "Colitis Gravis"[Title/Abstract] OR "Crohn Disease"[MeSH Terms] OR "Crohn Disease"[Title/Abstract] OR "Crohn's Enteritis"[Title/Abstract] OR "Regional Enteritis"[Title/Abstract] OR "Crohn's Disease"[Title/Abstract] OR "Crohns Disease"[Title/Abstract] OR "Granulomatous Enteritis"[Title/Abstract] OR "Ileocolitis"[Title/Abstract] OR "Granulomatous Colitis"[Title/Abstract] OR "Terminal Ileitis"[Title/Abstract] OR "Regional Ileitis"[Title/Abstract])  AND  ("Colorectal Neoplasms"[MeSH Terms] OR "Colorectal Neoplasms"[Title/Abstract] OR "colorectal neoplasm"[Title/Abstract] OR "colorectal neoplasms"[Title/Abstract] OR "Colorectal Cancer"[Title/Abstract]  OR "colorectal carcinoma"[Title/Abstract] OR "Colon Neoplasms"[MeSH Terms] OR "Colon Neoplasms"[Title/Abstract] OR "colon neoplasm"[Title/Abstract] OR "colon neoplasms"[Title/Abstract]  OR "Colon Cancer"[Title/Abstract] OR "colon carcinoma"[Title/Abstract] OR "Large Intestine Neoplasms"[MeSH Terms] OR "Large Intestine Neoplasms"[Title/Abstract] OR "large intestine neoplasm"[Title/Abstract] OR "large intestine neoplasms"[Title/Abstract] OR "Bowel Neoplasms"[MeSH Terms] OR "Bowel Neoplasms"[Title/Abstract] OR "bowel neoplasm"[Title/Abstract] OR "bowel neoplasms"[Title/Abstract] OR "Bowel Cancer"[Title/Abstract] OR "bowel carcinoma"[Title/Abstract]  OR "Rectal Neoplasms"[MeSH Terms] OR "Rectal Neoplasms"[Title/Abstract] OR "rectal neoplasm"[Title/Abstract] OR "rectal neoplasms"[Title/Abstract] OR "Rectal Cancer"[Title/Abstract] OR "rectal carcinoma"[Title/Abstract] OR "Colonic Neoplasms"[MeSH Terms] OR "Colonic Neoplasms"[Title/Abstract] OR "colonic neoplasm"[Title/Abstract] OR "colonic neoplasms"[Title/Abstract] OR "Rectosigmoid Neoplasms"[MeSH Terms] OR "Rectosigmoid Neoplasms"[Title/Abstract] OR "rectosigmoid neoplasm"[Title/Abstract] OR "rectosigmoid neoplasms"[Title/Abstract]) |

**Supplementary Appendix 2 Python Codes for Data and Analysis**

```
import matplotlib.pyplot as plt
import numpy as np
import pandas as pd
from matplotlib.patches import Polygon

# ──────────────────────────────────────────────
# DATASETS
# ──────────────────────────────────────────────
upf_ibd = pd.DataFrame({
    "Study": [
        "Narula 2021 – any IBD",
        "Lo 2022 – Crohn’s",
        "Lo 2022 – UC",
        "UK Biobank SSB – IBD",
        "EPIC 2023 – Crohn’s",
        "EPIC 2023 – UC",
        "Lifelines 2022 – Western (CD)",
        "Lifelines 2022 – Carnivorous (UC)",
    ],
    "HR":    [1.82, 1.70, 1.08, 1.94, 1.48, 0.93, 1.16, 1.11],
    "Lower": [1.22, 1.23, 0.78, 1.52, 0.79, 0.61, 1.03, 1.02],
    "Upper": [2.72, 2.35, 1.50, 2.49, 2.76, 1.43, 1.30, 1.21],
})

ibd_crc = pd.DataFrame({
    "Study": [
        "Olén 2020 – UC",
        "Olén 2020 – Crohn’s",
        "Jess 2012 – UC",
        "Jess 2012 – Crohn’s",
    ],
    "HR":    [1.66, 1.40, 1.07, 0.85],
    "Lower": [1.57, 1.27, 0.95, 0.67],
    "Upper": [1.76, 1.53, 1.21, 1.07],
})

upf_crc = pd.DataFrame({
    "Study": ["Wang 2022 – Men", "Wang 2022 – Women", "Chang 2022 – UK Biobank"],
    "HR":    [1.29, 1.04, 1.01],
    "Lower": [1.08, 0.90, 0.42],
    "Upper": [1.53, 1.20, 1.41],
})

# ──────────────────────────────────────────────
# FOREST PLOT FUNCTION (DerSimonian–Laird RE)
# ──────────────────────────────────────────────
def forest_random(df, title, output_path,
                  xlim=(0.4, 3.0),
                  xticks=(0.4, 0.5, 0.75, 1, 1.5, 2, 3)):
    """
    Draws a log-scale forest plot with DerSimonian–Laird random-effects pooling.
    """
    log_hr = np.log(df["HR"])
    se     = (np.log(df["Upper"]) - np.log(df["Lower"])) / (2 * 1.96)
    var    = se**2

    # Fixed-effect weights for Q statistic
    w_fe      = 1 / var
    pooled_fe = np.sum(w_fe * log_hr) / np.sum(w_fe)
    Q         = np.sum(w_fe * (log_hr - pooled_fe)**2)
    df_q      = len(df) - 1

    # DerSimonian–Laird tau-squared
    C    = np.sum(w_fe) - (np.sum(w_fe**2) / np.sum(w_fe))
    tau2 = max((Q - df_q) / C, 0)

    # Random-effects weights
    w_re = 1 / (var + tau2)

    # Pooled random-effects estimate
    re_log_hr    = np.sum(w_re * log_hr) / np.sum(w_re)
    re_se        = np.sqrt(1 / np.sum(w_re))
    re_hr        = np.exp(re_log_hr)
    re_ci_lower  = np.exp(re_log_hr - 1.96 * re_se)
    re_ci_upper  = np.exp(re_log_hr + 1.96 * re_se)

    # Plot
    fig, ax = plt.subplots(figsize=(7, 0.6 * len(df) + 2))
    y_positions = np.arange(len(df), 0, -1)

    ax.errorbar(
        df["HR"], y_positions,
        xerr=[df["HR"] - df["Lower"], df["Upper"] - df["HR"]],
        fmt='o', capsize=3, label='Study estimates'
    )

    # Pooled diamond
    diamond = Polygon([
        (re_ci_lower, 0),
        (re_hr,       0.3),
        (re_ci_upper, 0),
        (re_hr,      -0.3),
    ], closed=True, facecolor='lightgray', edgecolor='black', label='Pooled (RE)')
    ax.add_patch(diamond)

    # Formatting
    ax.set_yticks(list(y_positions) + [0])
    ax.set_yticklabels(list(df["Study"]) + ["Pooled"])
    ax.set_xscale('log')
    ax.set_xlim(xlim)
    ax.set_xticks(xticks)
    ax.axvline(1, linestyle='--', color='gray')
    ax.set_xlabel('Hazard Ratio (log scale)')
    ax.set_title(title, fontsize=12)
    ax.set_ylim(-1, len(df) + 1)

    plt.tight_layout()
    fig.savefig(output_path)
    plt.close(fig)

# ──────────────────────────────────────────────
# FUNNEL PLOT FUNCTION
# ──────────────────────────────────────────────
def funnel_plot(df, title, output_path):
    df = df.copy()
    df["logHR"] = np.log(df["HR"])
    df["SE"]    = (np.log(df["Upper"]) - np.log(df["Lower"])) / (2 * 1.96)

    weights      = 1 / df["SE"]**2
    pooled_loghr = np.sum(weights * df["logHR"]) / np.sum(weights)

    se_max       = df["SE"].max()
    se_vals      = np.linspace(0, se_max, 200)
    delta        = 1.96 * se_vals

    fig, ax = plt.subplots(figsize=(5, 5))
    ax.scatter(df["logHR"], df["SE"], s=30)
    ax.axvline(pooled_loghr, linestyle='--', color='gray')
    ax.plot(pooled_loghr + delta, se_vals, linestyle='--', color='gray')
    ax.plot(pooled_loghr - delta, se_vals, linestyle='--', color='gray')

    ax.set_xlabel('log(HR)')
    ax.set_ylabel('Standard Error')
    ax.set_title(title)
    ax.set_ylim(se_max + se_max * 0.1, 0)

    plt.tight_layout()
    fig.savefig(output_path)
    plt.close(fig)
```

| **Section and Topic** | **Item #** | **Checklist item** | **Location where item is reported** |
| --- | --- | --- | --- |
| **TITLE** | | |  |
| Title | 1 | Identify the report as a systematic review. | Page 1 |
| **ABSTRACT** | | |  |
| Abstract | 2 | See the PRISMA 2020 for Abstracts checklist. | Page 1 |
| **INTRODUCTION** | | |  |
| Rationale | 3 | Describe the rationale for the review in the context of existing knowledge. | Page 2 |
| Objectives | 4 | Provide an explicit statement of the objective(s) or question(s) the review addresses. | Page 2 |
| **METHODS** | | |  |
| Eligibility criteria | 5 | Specify the inclusion and exclusion criteria for the review and how studies were grouped for the syntheses. | Page 3 |
| Information sources | 6 | Specify all databases, registers, websites, organisations, reference lists and other sources searched or consulted to identify studies. Specify the date when each source was last searched or consulted. | Page 3 |
| Search strategy | 7 | Present the full search strategies for all databases, registers and websites, including any filters and limits used. | Page 3 |
| Selection process | 8 | Specify the methods used to decide whether a study met the inclusion criteria of the review, including how many reviewers screened each record and each report retrieved, whether they worked independently, and if applicable, details of automation tools used in the process. | Page 4 |
| Data collection process | 9 | Specify the methods used to collect data from reports, including how many reviewers collected data from each report, whether they worked independently, any processes for obtaining or confirming data from study investigators, and if applicable, details of automation tools used in the process. | Page 6 |
| Data items | 10a | List and define all outcomes for which data were sought. Specify whether all results that were compatible with each outcome domain in each study were sought (e.g. for all measures, time points, analyses), and if not, the methods used to decide which results to collect. | Page 6 |
|  | 10b | List and define all other variables for which data were sought (e.g. participant and intervention characteristics, funding sources). Describe any assumptions made about any missing or unclear information. | Page 6 |
| Study risk of bias assessment | 11 | Specify the methods used to assess risk of bias in the included studies, including details of the tool(s) used, how many reviewers assessed each study and whether they worked independently, and if applicable, details of automation tools used in the process. | Page 6 |
| Effect measures | 12 | Specify for each outcome the effect measure(s) (e.g. risk ratio, mean difference) used in the synthesis or presentation of results. | Page 8 |
| Synthesis methods | 13a | Describe the processes used to decide which studies were eligible for each synthesis (e.g. tabulating the study intervention characteristics and comparing against the planned groups for each synthesis (item #5)). | Page 8 |
|  | 13b | Describe any methods required to prepare the data for presentation or synthesis, such as handling of missing summary statistics, or data conversions. | Page 8 |
|  | 13c | Describe any methods used to tabulate or visually display results of individual studies and syntheses. | Page 8 |
|  | 13d | Describe any methods used to synthesize results and provide a rationale for the choice(s). If meta-analysis was performed, describe the model(s), method(s) to identify the presence and extent of statistical heterogeneity, and software package(s) used. | Page 8 |
|  | 13e | Describe any methods used to explore possible causes of heterogeneity among study results (e.g. subgroup analysis, meta-regression). | Page 8 |
|  | 13f | Describe any sensitivity analyses conducted to assess robustness of the synthesized results. | Page 8 |
| Reporting bias assessment | 14 | Describe any methods used to assess risk of bias due to missing results in a synthesis (arising from reporting biases). | Page 7 |
| Certainty assessment | 15 | Describe any methods used to assess certainty (or confidence) in the body of evidence for an outcome. | Page 13 |
| **RESULTS** | | |  |
| Study selection | 16a | Describe the results of the search and selection process, from the number of records identified in the search to the number of studies included in the review, ideally using a flow diagram. | Page 10 |
|  | 16b | Cite studies that might appear to meet the inclusion criteria, but which were excluded, and explain why they were excluded. | Page 10 |
| Study characteristics | 17 | Cite each included study and present its characteristics. | Page 11 |
| Risk of bias in studies | 18 | Present assessments of risk of bias for each included study. | Page 7 |
| Results of individual studies | 19 | For all outcomes, present, for each study: (a) summary statistics for each group (where appropriate) and (b) an effect estimate and its precision (e.g. confidence/credible interval), ideally using structured tables or plots. | Page 11 |
| Results of syntheses | 20a | For each synthesis, briefly summarise the characteristics and risk of bias among contributing studies. | Page 12 |
|  | 20b | Present results of all statistical syntheses conducted. If meta-analysis was done, present for each the summary estimate and its precision (e.g. confidence/credible interval) and measures of statistical heterogeneity. If comparing groups, describe the direction of the effect. | Page 12 |
|  | 20c | Present results of all investigations of possible causes of heterogeneity among study results. | Page 15 |
|  | 20d | Present results of all sensitivity analyses conducted to assess the robustness of the synthesized results. | Page 15 |
| Reporting biases | 21 | Present assessments of risk of bias due to missing results (arising from reporting biases) for each synthesis assessed. | Page 15 |
| Certainty of evidence | 22 | Present assessments of certainty (or confidence) in the body of evidence for each outcome assessed. | Page 13 |
| **DISCUSSION** | | |  |
| Discussion | 23a | Provide a general interpretation of the results in the context of other evidence. | Page 16 |
|  | 23b | Discuss any limitations of the evidence included in the review. | Page 16 |
|  | 23c | Discuss any limitations of the review processes used. | Page 16 |
|  | 23d | Discuss implications of the results for practice, policy, and future research. | Page 16 |
| **OTHER INFORMATION** | | |  |
| Registration and protocol | 24a | Provide registration information for the review, including register name and registration number, or state that the review was not registered. | Page 3 |
|  | 24b | Indicate where the review protocol can be accessed, or state that a protocol was not prepared. | Page 3 |
|  | 24c | Describe and explain any amendments to information provided at registration or in the protocol. | Page 3 |
| Support | 25 | Describe sources of financial or non-financial support for the review, and the role of the funders or sponsors in the review. | Page 17 |
| Competing interests | 26 | Declare any competing interests of review authors. | Page 17 |
| Availability of data, code and other materials | 27 | Report which of the following are publicly available and where they can be found: template data collection forms; data extracted from included studies; data used for all analyses; analytic code; any other materials used in the review. | Page 17 |

*From:*  Page MJ, McKenzie JE, Bossuyt PM, Boutron I, Hoffmann TC, Mulrow CD, et al. The PRISMA 2020 statement: an updated guideline for reporting systematic reviews. BMJ 2021;372:n71. doi: 10.1136/bmj.n71

For more information, visit: <http://www.prisma-statement.org/>
